## Supplement for "Accuracy of digital chest x-ray analysis with artificial intelligence software as a triage and screening tool in hospitalized patients being evaluated for tuberculosis in Lima, Peru"

*qXR version 3*

Using the culture reference standard for pulmonary tuberculosis, qXR v3 (at the manufacturer pre-specified threshold of 0.5) had an overall sensitivity of 0.91 (59/65, 95% CI 0.81-0.97), specificity of 0.32 (102/322, 95% CI 0.27-0.37), and AUC of 0.78 (95% CI 0.72-0.84) (Table S5). Using the Xpert reference standard for pulmonary tuberculosis, qXR v3 (at the manufacturer pre-specified threshold of 0.5) had an overall sensitivity of 0.93 (64/69, 95% CI 0.84-0.98), specificity of 0.32 (105/329, 95% CI 0.31-0.41), and AUC of 0.76 (95% CI 0.70-0.82) (Table S5). When sensitivity was set at 90% to match the WHO triage test accuracy performance criteria, specificity rose to 0.41 (133/320, 95% CI 0.36-0.47) and 0.37 (121/329, 95% CI 0.32-0.42) with the culture and Xpert reference standards respectively (Table S5).

Stratified analyses

There was no difference in qXR v3 sensitivity when stratified by sex, age, prior TB, HIV, symptoms, or smear status (Figure S1). qXR v3 specificity was higher in people without prior TB, people with cough less than 2 weeks, and who did not report weight loss (Figure S2).

Screening cohort

Since there was only one person with confirmed TB in the screening group (who did have a qXR positive result), we only report specificity. Using the manufacturer’s pre-specified thresholds, the specificity for qXR v3 was 0.93 (95% CI 0.89-0.97) using the culture reference standard and 0.96 (95% CI 0.92-0.98) using the Xpert reference standard (Table S3).

Table S1a: Summary of Culture and Xpert result concordance.

|  | Culture Negative  (n=322) | Culture  Positive  (n=65) | Missing  Culture  (n=52) |
| --- | --- | --- | --- |
| Xpert Negative  (n=329) | 298 | 4 | 27 |
| Xpert Positive  (n=69) | 9 | 55 | 5 |
| Missing Xpert  (n=41) | 15 | 6 | 20 |

Table S1b: Xpert sensitivity for smear-positive and smear-negative culture-positive TB in triage cohort patients

|  | Sensitivity – smear positive culture positive  (95% CI) | Sensitivity – smear negative  culture positive  (95% CI) |
| --- | --- | --- |
| Xpert | (35/37)  94.6%  (81.8-99.3%) | (12/14)  85.7%  (57.2-98.2%) |

Table S2: Demographic and clinical characteristics of enrolled participants compared to those who were excluded.

|  | Triage (n=419) | Screening (n=184) | Excluded-  Triage (n=475) | P-value* | Excluded- Screening  (n=24) | P-value** |
| --- | --- | --- | --- | --- | --- | --- |
| **Median Age (years, interquartile range)** | 41.35 (26.8, 56.6) | 36.19 (25.19, 50.53) | 37.33 (26.2, 51.8) | **0.028** | 35.27 (24.7, 45.6) | 0.632 |
| **Sex, No (%)**  Female  Male | 164 (39.1)  255 (60.9) | 111 (60.3)  73 (39.7) | 246 (51.8)  229 (48.2) | **<0.001** | 14 (58.3)  10 (41.7) | 1.000 |
| **History of Previous TB, No (%)**  Yes  No  Refused | 140 (33.4)  278 (66.4)  1 (0.20) | 0 (0.00)  184 (100)  0 (0.00) | 158 (33.3)  317 (66.7)  0 (0.00) | 0.771 | 0 (0.00)  24 (100)  0 (0.00) | - |
| **HIV, No (%)**  Yes  No | 36 (8.6)  383 (91.4) | 1 (0.5)  183 (99.5) | 29 (6.1)  446 (93.9) | 0.158 | 1 (4.2)  23 (95.8) | 0.218 |
| **Smoking, No (%)**  Never  Former  Current | 202 (48.2)  161 (38.4)  56 (13.4) | 99 (53.8)  51 (27.7)  34 (18.5) | 292 (61.5)  133 (28.0)  50 (10.5) | **<0.001** | 13 (54.2)  6 (25.0)  5 (20.8) | 0.944 |
| **Alcohol, No (%)**  Never  Former  Current  Missing | 107 (25.5)  121 (28.9)  189 (45.1)  2 (0.5) | 32 (17.3)  33 (18.0)  119 (64.7)  0 (0.00) | 125 (26.3)  105 (22.1)  244 (51.4)  1 (0.2) | 0.242 | 4 (16.7)  7 (29.2)  13 (54.2)  0 (0.00) | 0.433 |
| **Respiratory Disease, No (%)**  Asthma  Bronchiectasis  None | 28 (6.7)  13 (3.1)  378 (90.2) | 2 (1.1)  0 (0.00)  182 (98.9) | 22 (4.6)  17 (3.6)  436 (91.8) | 0.390 | 0 (0.00)  0 (0.00)  24 (100) | 0.608 |
| **Diabetes, Type II, No (%)**  Yes  No | 58 (13.8)  361 (86.2) | 25 (13.6)  159 (86.4) | 44 (9.3)  431 (90.7) | **0.035** | 2 (8.3)  22 (91.7) | 0.747 |
| **Prison, No (%)**  Yes  No | 62 (14.8)  357 (85.2) | 3 (1.6)  181 (98.4) | 41 (8.6)  434 (91.4) | **0.005** | 1 (4.2)  23 (95.8) | 0.390 |
| **Household Contact of TB positive patient, No (%)**  Yes  No  Missing | 159 (38.0)  256 (61.1)  4 (0.9) | - | 152 (32.0)  317 (66.7)  6 (1.3) | 0.165 | 3 (12.5)  21 (87.5)  0 (0.00) | - |
| **Smear Status, No (%)**  Positive  Negative  Missing | 48 (11.5)  363 (86.6)  8 (1.9) | 0 (0.00)  183 (99.5)  1 (0.5) | 54 (11.4)  411 (86.5)  11 (2.3) | 0.420 | 0 (0.00)  19 (79.2)  5 (20.8) | 1.000 |
| **Cough, No (%)**  *Length, in Weeks*  Less than 1 week  1-2 weeks  More than 2 weeks  Missing  *Phlegm*  Yes  No  Missing  *Blood*  Yes  No  Missing | 102 (24.4)  107 (25.5)  189 (45.1)  21 (5.0)  352 (84.0)  47 (11.2)  20 (4.8)  166 (39.6)  233 (55.6)  20 (4.8) | -  -  - | 148 (31.2)  97 (20.4)  189 (39.8)  41 (8.6)  373 (78.5)  64 (13.5)  38 (8.0)  135 (28.4)  302 (63.6)  38 (8.0) | 0.065  0.071  **0.001** | -  -  - | -  -  - |
| **Fever, No (%)**  Yes  No  Refused | 265 (63.3)  153 (36.5)  1 (0.2) | 85 (46.2)  99 (53.8)  0 (0) | 248 (52.2)  227 (47.8)  0 (0.0) | **0.001** | 7 (29.2)  17 (70.8)  0 (0.00) | 0.130 |
| **Night Sweats in the last 3 months, No (%)**  Yes  No  Refused | 251 (59.9)  168 (40.1)  0 (0.0) | 52 (28.3)  132 (71.7)  0 (0.0) | 248 (52.2)  226 (47.6)  1 (0.2) | **0.026** | 5 (20.8)  19 (79.2)  0 (0.0) | 0.627 |
| **Weight Loss (unintentional), No (%)**  Yes  No  Refused | 293 (69.9)  123 (29.4)  3 (0.7) | 84 (45.6)  98 (53.3)  2 (1.1) | 293 (61.7)  182 (38.3)  0 (0.0) | **0.002** | 10 (41.6)  13 (54.2)  1 (4.2) | 0.349 |
| **Difficulty Breathing, No (%)**  Yes  No | 335 (80.0)  84 (20.0) | 52 (28.3)  132 (71.7) | 310 (65.3)  165 (34.7) | **<0.001** | 6 (25.0)  18 (75.0) | 0.814 |

*Triage vs Excluded from triage cohort

**Screening vs Excluded from screening cohort

Table S3: Diagnostic accuracy of qXR Version 3 and 4 using a reference standard which is positive if either mycobacterial culture or Xpert is positive for the triage cohort.

|  | Sensitivity  (95% CI) | Specificity  (95% CI) | AUC  (95% CI) |
| --- | --- | --- | --- |
| qXR Version 3 | | | |
| Manufacturer Threshold 0.5 | (73/79)  92.4%  (84.2-97.2%) | (96/298)  32.2%  (26.9-37.8%) | 0.778  (0.719-0.837) |
| qXR Version 4 | | | |
| Manufacturer Threshold 0.5 | (73/79)  92.4%  (84.2-97.2%) | (97/298)  32.6%  (27.3-38.2%) | 0.775  (0.716-0.834) |

Table S4: Diagnostic accuracy of qXR Version 3 and 4 for pre-specified subgroups in the triage cohort for which participants with prior TB, respiratory diseases, and HIV, were excluded

|  | Excluding Patients with Prior TB | | | Excluding Patients with Respiratory Diseases | | | Excluding People with HIV | | |
| --- | --- | --- | --- | --- | --- | --- | --- | --- | --- |
| Threshold | Sensitivity  (95% CI) | Specificity  (95% CI) | AUC  (95% CI) | Sensitivity  (95% CI) | Specificity  (95% CI) | AUC  (95% CI) | Sensitivity  (95% CI) | Specificity  (95% CI) | AUC (95% CI) |
| Culture | | | | | | | | | |
| qXR Version 3 | | | | | | | | | |
| Manufacturer Threshold 0.5 | (46/52)  88.5% (76.6-95.6%) | (84/208)  40.4% (33.7-47.4%) | 0.800 (0.730, 0.870) | (58/64)  90.6% (80.7-96.5%) | (92/285)  32.3% (26.9-38.0%) | 0.784 (0.719, 0.848) | (54/59)  91.5% (81.3-97.2%) | (89/294)  30.3% (25.1-35.9%) | 0.792 (0.730, 0.854) |
| qXR Version 4 | | | | | | | | | |
| Manufacturer Threshold 0.5 | (46/52)  88.5% (76.6-95.6%) | (84/208)  40.4%  (33.7-47.4%) | 0.802 (0.731, 0.872) | (58/64)  90.6% (80.7-96.5%) | (93/285)  32.6% (27.2-38.4%) | 0.781 (0.716, 0.846) | (54/58)  93.1% (83.3-98.1%) | (90/294)  30.6%  (25.4-36.2%) | 0.789 (0.726, 0.851) |
| Xpert | | | | | | | | | |
| qXR Version 3 | | | | | | | | | |
| Manufacturer Threshold 0.5 | (49/54)  90.7%  (79.7-96.9%) | (87/213)  40.8% (34.2-47.8%) | 0.784 (0.715, 0.854) | (63/68)  92.6% (83.7-97.6%) | (94/292)  32.2% (26.9-37.9%) | 0.763 (0.698, 0.827) | (58/61)  95.1% (86.3-99%) | (92/302)  30.5% (25.3-36%) | 0.778 (0.716, 0.840) |
| qXR Version 4 | | | | | | | | | |
| Manufacturer Threshold 0.5 | (49/54)  90.7% (79.7-96.9%) | (86/213)  40.4%  (33.7-47.3%) | 0.784 (0.714, 0.854) | (63/68)  92.6% (83.7-97.6%) | (95/292)  32.5% (27.2-38.2%) | 0.758 (0.693, 0.823) | (58/61)  95.1% (86.3-99%) | (93/302)  30.8% (25.6-36.3%) | 0.775 (0.712, 0.837) |

Table S5: Summary of Diagnostic Accuracy for qXR version 3 compared to the culture (primary) and Xpert (secondary) reference standards

|  | Triage Patients | | | Screening Patients | | |
| --- | --- | --- | --- | --- | --- | --- |
|  | Sensitivity  (95% CI) | Specificity  (95% CI) | AUC  (95% CI) | Sensitivity  (95% CI) | Specificity  (95% CI) | AUC  (95% CI) |
| Culture | | | | | | |
| qXR Version 3 | | | | | | |
| Manufacturer Threshold 0.5 | 90.8%  59/65  (81-96.5%) | 31.7%  102/322  (26.6-37.1%) | 0.780  (0.717, 0.843) | ^ | 94.2%  162/172  (89.5-96.9%) | - |
| Threshold 0.675* | 90.8%  59/65  (81-96.5%) | 41.3%  133/320  (35.9-46.9%) | - | ^ | 97.1%  167/172  (93.2-98.8%) | - |
| Xpert | | | | | | |

| qXR Version 3 | | | | | | |
| --- | --- | --- | --- | --- | --- | --- |
| Manufacturer Threshold 0.5 | 92.8%  64/69  (83.9-97.6%) | 31.9%  105/329  (26.9-37.3%) | 0.759  (0.696, 0.821) | 100%  1/1  (2.5-100%) | 94.4%  169/179  (90.0-97.3%) | 1.00  (-, 1.00) |
| Threshold 0.6* | 89.9%  62/69  (80.2-95.8%) | 36.8%  121/329  (31.6-42.2%) | - | 100%  1/1  (2.5-100%) | 97.2%  174/179  (93.6-99.1%) | - |

*threshold at which sensitivity is closest to 90%

^No positive cultures in the Screening Group

Table S6: Lung abnormalities detected by qXR analysis for the Triage and Screening cohorts

|  | **Triage**  (N=419)  n/N, %  (95% CI) | **Screening**  (N=184)  n/N, %  (95% CI) |
| --- | --- | --- |
| Atelectasis | 93/419  22.2%  (18.4-26.4%) | 14/184  7.61%  (4.5-12.5%) |
| Blunted CP | 17/419  4.06%  (2.5-6.4%) | 5/184  2.72%  (1.1-6.4%) |
| Calcification | 68/419  16.2%  (13.0-20.1%) | 2/184  1.09%  (0.3-4.3%) |
| Cardiomegaly | 36/419  8.59%  (6.3-11.7%) | 17/184  9.24%  (5.8-14.4%) |
| Cavity | 80/419  19.1%  (15.6-23.2%) | 0/184  0%  (0-0%) |
| Consolidation | 261/419  62.3%  (57.5-66.8%) | 7/184  3.80%  (1.8-7.8%) |
| Fibrosis | 195/419  46.5%  (41.8-51.3%) | 19/184  10.3%  (6.7-15.7%) |
| Hilar Lymphadenopathy | 2/419  0.48%  (0.1-1.9%) | 0/184  0%  (0-0%) |
| Nodule | 242/419  57.8%  (53.0-62.4%) | 20/184  10.9%  (7.1-16.3%) |
| Opacity | 341/419  81.4%  (77.4-84.8%) | 44/184  23.9%  (18.3-30.7%) |
| Pleural Effusion | 151/419  36.0%  (31.6-40.8%) | 9/184  4.89%  (2.6-9.2%) |

### Reporting checklist for diagnostic test accuracy study.

Based on the STARD guidelines.

|  |  | Reporting Item | Page Number |
| --- | --- | --- | --- |
| **Title or abstract** |  |  | 1 |
| None | [#1](https://www.goodreports.org/reporting-checklists/stard/info/#1) | Identification as a study of diagnostic accuracy using at least one measure of accuracy (such as sensitivity, specificity, predictive values, or AUC) |  |
| 1 |  |  |  |
| **Abstract** |  |  | 1-2 |
| None | [#2](https://www.goodreports.org/reporting-checklists/stard/info/#2) | Structured summary of study design, methods, results, and conclusions (for specific guidance, see STARD for Abstracts https://www.equator-network.org/reporting-guidelines/stard-abstracts/) |  |
| 1-2 |  |  |  |
| **Introduction** |  |  |  |
| None | [#3](https://www.goodreports.org/reporting-checklists/stard/info/#3) | Scientific and clinical background, including the intended use and clinical role of the index test | 3 |
| None | [#4](https://www.goodreports.org/reporting-checklists/stard/info/#4) | Study objectives and hypotheses | 4-5 |
| **Methods** |  |  |  |
| Study design | [#5](https://www.goodreports.org/reporting-checklists/stard/info/#5) | Whether data collection was planned before the index test and reference standard were performed (prospective study) or after (retrospective study) |  |
| 5 |  |  |  |
| Participants | [#6](https://www.goodreports.org/reporting-checklists/stard/info/#6) | Eligibility criteria | 5 |
| Participants | [#7](https://www.goodreports.org/reporting-checklists/stard/info/#7) | On what basis potentially eligible participants were identified (such as symptoms, results from previous tests, inclusion in registry) |  |
| 5 |  |  |  |
| Participants | [#8](https://www.goodreports.org/reporting-checklists/stard/info/#8) | Where and when potentially eligible participants were identified (setting, location and dates) | 5 |
| Participants | [#9](https://www.goodreports.org/reporting-checklists/stard/info/#9) | Whether participants formed a consecutive, random or convenience series | 5-6 |
| Test methods | [#10](https://www.goodreports.org/reporting-checklists/stard/info/#10) | Index and reference tests in sufficient detail to allow replication | 5-6 |
| Test methods | [#11](https://www.goodreports.org/reporting-checklists/stard/info/#11) | Rationale for choosing the reference standard (if alternatives exist) | 6-7 |
| Test methods | [#12](https://www.goodreports.org/reporting-checklists/stard/info/#12) | Definition of and rationale for test positivity cut-offs or result categories of the index and reference tests, distinguishing pre-specified from exploratory |  |
| 6-7 |  |  |  |
| Test methods | [#13](https://www.goodreports.org/reporting-checklists/stard/info/#13) | Whether clinical information and reference standard results were available to the performers / readers of the index test; Whether clinical information and index test results were available to the assessors of the reference standard |  |
| 7-8 |  |  |  |
| Analysis | [#14](https://www.goodreports.org/reporting-checklists/stard/info/#14) | Methods for estimating or comparing measures of diagnostic accuracy | 7 |
| Analysis | [#15](https://www.goodreports.org/reporting-checklists/stard/info/#15) | How indeterminate index test or reference standard results were handled | 7 |
| Analysis | [#16](https://www.goodreports.org/reporting-checklists/stard/info/#16) | How missing data on the index test and reference standard were handled | 7 |
| Analysis | [#17](https://www.goodreports.org/reporting-checklists/stard/info/#17) | Any analyses of variability in diagnostic accuracy, distinguishing pre-specified from exploratory | 7 |
| Analysis | [#18](https://www.goodreports.org/reporting-checklists/stard/info/#18) | Intended sample size and how it was determined | 7 |
| **Results** |  |  |  |
| Participants | [#19](https://www.goodreports.org/reporting-checklists/stard/info/#19) | Flow of participants, using a diagram | 8 |
| Participants | [#20](https://www.goodreports.org/reporting-checklists/stard/info/#20) | Baseline demographic and clinical characteristics of participants | 8-10 |
| Participants | [#21](https://www.goodreports.org/reporting-checklists/stard/info/#21) | Distribution of severity of disease in those with the target condition, and distribution of alternative diagnoses in those without the target condition | 8-10 |
| Participants | [#22](https://www.goodreports.org/reporting-checklists/stard/info/#22) | Time interval and any clinical interventions between index test and reference standard | 8-9 |
| Test results | [#23](https://www.goodreports.org/reporting-checklists/stard/info/#23) | Cross tabulation of the index test results (or their distribution) by the results of the reference standard |  |
| 8 |  |  |  |
| Test results | [#24](https://www.goodreports.org/reporting-checklists/stard/info/#24) | Estimates of diagnostic accuracy and their precision (such as 95% confidence intervals) | 11-12 |
| Test results | [#25](https://www.goodreports.org/reporting-checklists/stard/info/#25) | Any adverse events from performing the index test or the reference standard | N/A |
| **Discussion** |  |  |  |
| None | [#26](https://www.goodreports.org/reporting-checklists/stard/info/#26) | Study limitations, including sources of potential bias, statistical uncertainty, and generalisability | 17 |
| None | [#27](https://www.goodreports.org/reporting-checklists/stard/info/#27) | Implications for practice, including the intended use and clinical role of the index test | 14-16 |
| **Other information** |  |  |  |
| None | [#28](https://www.goodreports.org/reporting-checklists/stard/info/#28) | Registration number and name of registry | 5 |
| None | [#29](https://www.goodreports.org/reporting-checklists/stard/info/#29) | Where the full study protocol can be accessed | 5 |
| None | [#30](https://www.goodreports.org/reporting-checklists/stard/info/#30) | Sources of funding and other support; role of funders | 19 |

The STARD checklist is distributed under the terms of the Creative Commons Attribution License CC-BY. This checklist was completed on 15. May 2023 using <https://www.goodreports.org/>, a tool made by the [EQUATOR Network](https://www.equator-network.org) in collaboration with [Penelope.ai](https://www.penelope.ai)
