## Supplementary figures and images for "Accuracy of digital chest x-ray analysis with artificial intelligence software as a triage and screening tool in hospitalized patients being evaluated for tuberculosis in Lima, Peru"

### Supplemental Figure 1

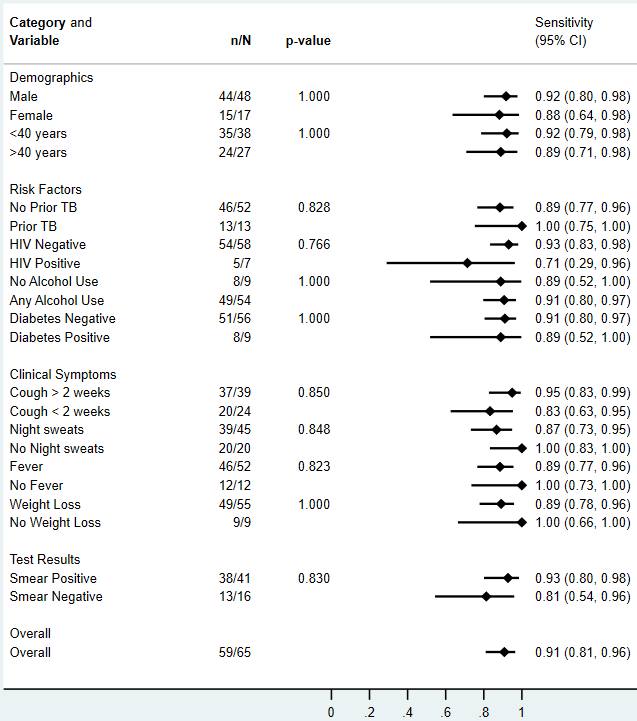

### Supplemental Figure 2

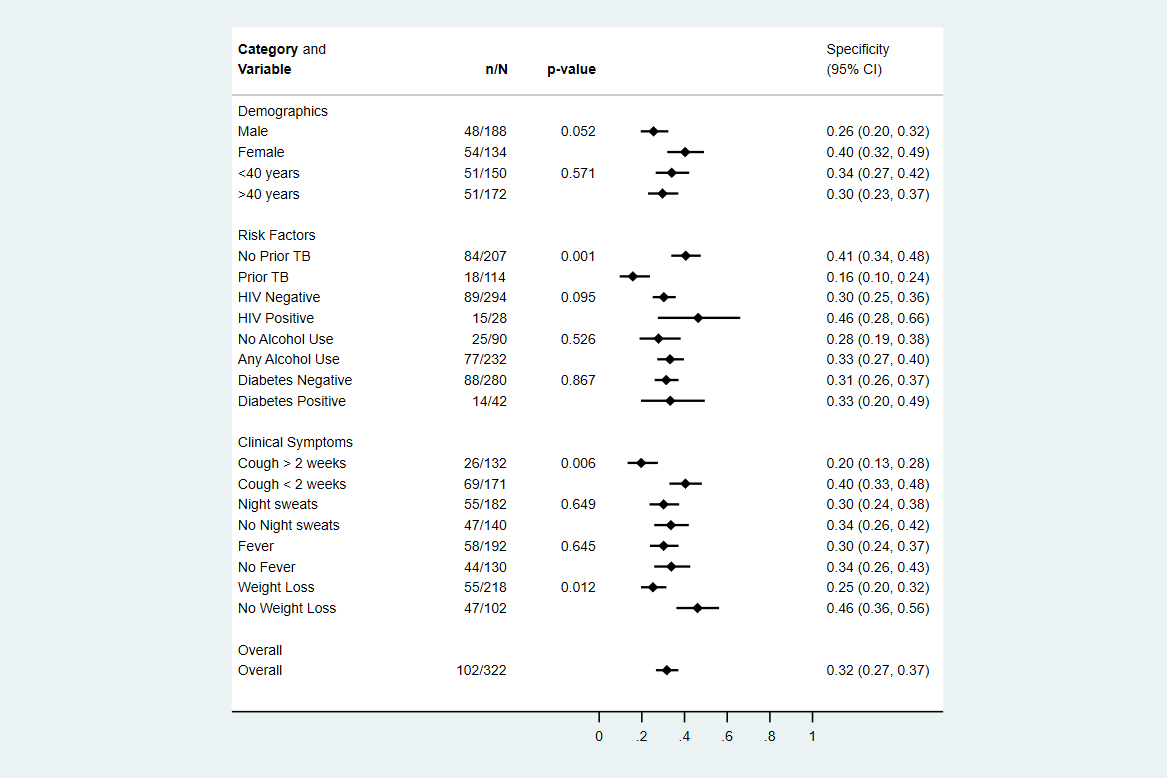
